# A machine learning approach for automating review of a RxNorm medication mapping pipeline output

**DOI:** 10.1101/2025.04.07.25325343

**Authors:** Matthias Hüser, John Doole, Vinicius Pinho, Hossein Rouhizadeh, Douglas Teodoro, Ahson Saiyed, Matvey B. Palchuk

## Abstract

Medication mapping to standardized terminologies is an important prerequisite for performing analytics on a federated EHR network. TriNetX LLC operates the largest such network in the world. Here we report on a novel pipeline, called `RxEmbed`, for the mapping and binding of local medication descriptions to RxNorm ingredient codes, using LLMs, and automated mapping review using machine learning. Performance of `RxEmbed` was assessed in a public data set from France as well as 6 Healthcare Organizations from the TriNetX federated EHR network across the United States and Brazil. On the public data set, `RxEmbed` outperformed two recently reported LLM-based baselines in terms of recall, and precision of generated mappings. In TriNetX network data, `RxEmbed` obtained RxNorm mapping recalls of 84-93 %, at a precision of 99.5-100 %. We built and evaluated a LLM-based medication mapping pipeline, that binds local medication descriptions from EHR systems to RxNorm ingredient codes. The high precision of the pipeline output implies very limited need for human review of the generated mappings.

## 1 Introduction

### Background

In the context of a federated electronic health record (EHR)-derived clinical data network, such as the one operated by TriNetX LLC [11, 16], it is of key importance to harmonize medication information to a common standard, so that data can be queried for cohort identification [20] and research. Faced with a large number of sites that are added to the network continuously, it is desirable to partially automate the task of code mapping, such as medications to a standard terminology using their descriptions. Beyond this specific need at TriNetX, the task of harmonizing medication data to a common standard is encountered in a variety of settings in the community. With the advent of large language models, such as GPT4o [2] or Llama [17], it is an open question how to leverage them for automating the mapping of medications, and the review of such mappings.

### Objective

We address the task of mapping local medication codes and associated descriptions to RxNorm [10], focusing specifically on ingredient codes. When codes are already delivered in a standard commercial or national terminology, mapping by code is preferable, and subsequently re-using the resulting mapping across the sites using the same terminology. However, mapping by code is not always possible as many sites use local coding systems. In these cases, we focused on the task of *mapping by description*, where a Natural Language Processing (NLP) approach–such as the one proposed here–is required. A key challenge is the need for extensive manual review that is typically required after generating candidate mappings [22].

## 2 Related work

Medical concept extraction and normalization (MCN) [20] is a classical task in the field of NLP for clinical content. For generating RxNorm bindings, most proposed toolkits use regular expression and dictionary matching approaches [8, 15, 18, 19]. In a recent MCN challenge, combinations of dictionary matching, word embeddings, cosine distance, and retrieve-and-rank techniques [4, 7] were used. Large language models have been previously used independently to assign standard codes to clinical narratives in German [1] or related tasks such as clinical phenotyping [6]. To our knowledge, a pipeline for mapping descriptions specifically to RxNorm ingredient codes with both LLM and machine learning components has not been reported in the literature, although some more general frameworks recently appeared [9]. In Table 1 we compare `RxEmbed` with other common techniques used in medication mapping pipelines. Recent studies found that the use of LLMs for MCN is not as straightforward as expected and faces several limitations. One example is fragility to substitutions of generics with brand names, when the model has to recognize brand and generic drug names as equivalent [5]. In a recent work a high sensitivity of LLMs to surface-level text variations, such as minor wording and formatting change was shown [14].

**Table 1:** RxEmbed’s main pipeline components put into context with other techniques used in previously proposed medication mapping pipelines.

| Technique | Used in RxEmbed? | Notes |
| --- | --- | --- |
| Translation | Yes | Standard technique to pre-process data. |
| Named entity recognition | Yes | Often used as pre-processing in similar works. |
| Regular expression matching | No | Often fragile to input format, but often used. |
| Dictionary matching | Yes | Nearest neighbour indices are constructed of OpenAI embeddings, and searching in them. |
| Word embeddings | Yes | OpenAI embeddings of identified tokens and RxNorm concepts are used. |
| Retrieve-and-rank | No | No explicit re-ranker is used, however reviewing retrieved concepts with LLMs serves a similar function. |
| Pre-review of mappings with LLMs | Yes | Rarely used in prior works for this task. |
| Automated mapping review with ML | Yes | Rarely used as post-processing for this task. |

## 3 Methods

### Input and outputs of the pipeline

Local medication codes and the associated descriptions, provided by a healthcare organization (HCO), are given as input to the pipeline. Since the codes themselves carry no semantic meaning, the pipeline relies on the descriptions to generate candidate mappings to RxNorm ingredient codes. As pre-processing any descriptions which are empty or invalid, and cannot be mapped by description, are filtered out. The output of the pipeline is a set of proposed mappings of local codes to RxNorm ingredient codes. These mappings can then be used to harmonize medication data from the respective HCO and be included in the TriNetX platform.

### Partial translation of medication descriptions

A partial translation step is performed only if the descriptions are not in English, which is the canonical language for the pipeline. For partial translation, we use `GPT4o` with a custom prompt. Hereby the model is instructed to leave untranslatable tokens such as medication brand names or numeric specifications unmodified.

**Figure 1.**
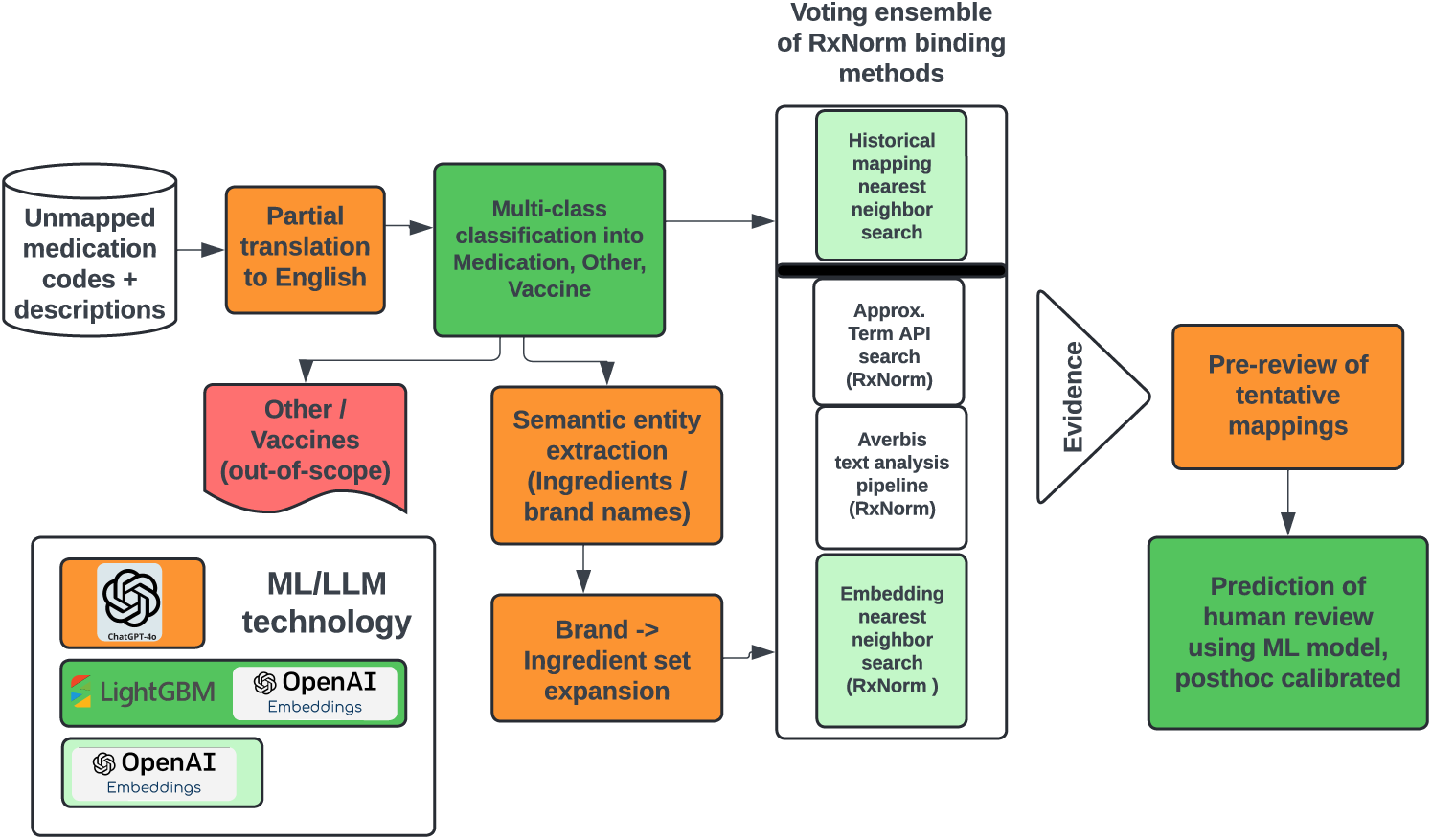
Overview of the components of the `RxEmbed` pipeline and the data flow through the pipeline from the local medication descriptions to the generated RxNorm mappings.

### Description category prediction

Based on our observation that a sizable proportion of the local descriptions do not actually refer to medications, but durable medical equipment, nutritional supplements or vaccines, we developed a multi-category classifier, that predicts the entity type denoted by the description. As machine learning features we use OpenAI `text-embedding-small` embeddings of the input descriptions (dimension 128). Data sets of local descriptions from TriNetX’s HCO network with examples of each category were hand-curated, providing the labels for machine learning. These data sets were derived through a different process than does not involve the evaluation data sets so no data leakage into the test set could occur. A gradient-boosting multi-category classifier is trained using `LightGBM` to place each new medication descriptions into one of the 5 classes:

- Strategic medication: A medication of key importance, such as drugs that appear frequently in cohort definitions, recently approved medications, and others.
- Other medication: All other medications.
- Fluid/Nutrition: Intravenous fluids and nutritional supplements and solutions, which have low relevance for patient cohort identification.
- Device/Procedure: Durable medical equipment or devices, potentially related to medication administration.
- Vaccines.

For the purpose of mapping to RxNorm, we focused on the first two categories. If a description is assigned to either ‘Fluid/Nutrition’, ‘Device/Procedure’ or ‘Vaccine’, these concepts are marked and handled via separate mechanism, outside of the scope of this pipeline.

### Semantic entity extraction

Medication entity extraction aims to extract entities from local descriptions which are candidates for binding to RxNorm ingredient codes. Separate prompts are passed to `GPT4o` and used to extract comma-separated lists of (1) ingredient tokens, as well as (2) medication brand name tokens from the input description.

### Expansion of brand name tokens to ingredients

As some medication descriptions might only contain brand names and lack explicit mentions of ingredients, in this step, a `GPT4o` prompt is used to expand brand name into ingredients. As context, we also pass the name of the country, from which the medication description originates as an argument to the prompt. To increase robustness and minimize LLM hallucinations, we use a second `GPT4o` prompt, that validates that predicted ingredients are indeed contained in a medication of the given brand. In some countries, hand-curated datasets are available that map medication brand names to their corresponding ingredients. For these countries, in addition to the `GPT4o`-based strategy, we created a nearest neighbour search index of brand names using OpenAI `text-embedding-small` embeddings, and when encountering a brand token, search in this index. If the retrieved brand name has a normalized Damerau-Levenshtein distance [3] of less than 0.5, the list of predicted ingredients is extended with this set.

### Binding of search terms to RxNorm ingredient codes

#### Voting ensemble approach

For binding of brand name tokens and extracted ingredients, which could be a token of the original string, or an ingredient predicted in the expansion step, we use four different sources of RxNorm mappings, which are combined in a voting approach: historical mappings, RxNorm’s Approximate Term API, Averbis text analysis pipeline, and nearest neighbor search using RxNorm embeddings.

#### Historical mappings

Historical mappings refers to the continuously evolving proprietary mappings database of local medication codes to RxNorm. Note that these mappings can be generated by this pipeline, but can also be generated by other means, such as mapping with code conversion tables supplied by the HCOs or using national terminology mappings and other methods. The descriptions of historical mappings are embedded using OpenAI `text-embedding-small` to vectors of dimension 128. We use the translated description to perform a nearest neighbor search in the set of historical mappings, on which a Open AI-based `text-embedding-small` index has been pre-computed. The five nearest neighbors are returned and accepted if the normalized Daverau-Levenshtein distance [3] of the description of the historical mapping and the search description is less than 0.5.

#### Approximate Term API

A local version of the Approximate Term API endpoint of RxNav [21] is used as a source for RxNorm code bindings. Extracted ingredient tokens and predicted ingredients are used as input `term`s to the API endpoint. Returned candidates are scanned in descending order of Approximate Term API match score, and the top candidate with term type of ingredient (`IN`), precise ingredient (`PIN`), or multi-ingredient (`MIN`) is used as a tentative match. `PIN` and `MIN` terms are subsequently normalized to `IN` terms. We compute the normalized Damerau-Levenshtein distance [3] between the target string, i.e., an ingredient description and the RxCUI description, and reject all tentative matches with a distance greater than 0.5.

#### Averbis text analysis pipeline

Another component is a proprietary text analysis pipeline provided by the Averbis (Averbis GmbH, Freiburg im Breisgau, Germany) Health Discovery NLP solution [12]. As input to the pipeline only the search term, either a brand name token, or an extracted ingredient is passed. We compute the normalized Damerau-Levenshtein distance [3] between the target string and the RxCUI description, and reject all tentative matches with a distance greater than 0.5.

#### RxNorm embedding and nearest neighbor search

A library of RxCUI ingredient code description embeddings is pre-computed with the OpenAI `text-embedding-small` model (embedding dimension of 128). For each input description string, a nearest neighbor search is performed to identify the closest neighbor in the index of embedded RxCUI descriptions. Like the previous method, the normalized Damerau-Levenshtein distance [3] between the target string and the RxCUI description is computed. Tentative matches with a distance of greater than 0.5 are rejected.

**Table 2:** Overview of GPT4o prompts and their mechanisms in the RxEmbed pipeline.

| Pipeline component | Function of prompt | Prompt details |
| --- | --- | --- |
| Partial translation | Translation | Translation should proceed word-wise and untranslatable tokens should be left unmodified. |
| Ingredient extraction | Semantic entity extraction | The agent should extract all substrings in the original string that describe active ingredients in a medication. |
| Brandname extraction | Semantic entity extraction | The agent should extract all substrings in the original string that describe brand-names of medications. |
| Ingredient prediction | Lookup of ingredients in medication brands | The agent should return the main active ingredients contained in a medication brand-name given as parameter the country where the brand is sold. |
| Ingredient review | Review lookup of ingredients in medication brands | This agent serves as a review agent for the ingredient prediction step and decides whether an ingredient is indeed contained in a brand, given the country as a parameter. |
| Ingredient mapping review | Review RxNorm ingredient code mappings | This agent serves as a review agent for any RxNorm ingredient code mappings that are generated by the pipeline. It is instructed to output an integer score between 0 and 10, reflecting its confidence that the mapping is correct. |

### Pre-review of ingredient mappings with GPT4o

Mappings proposed by at least one of the mapping sources in the voting ensemble are pre-reviewed by a `GPT4o` prompt to evaluate their medical plausibility. We pass the (potentially) translated local description of the medication, the description of generated RxCUI ingredient code, the country where the medication observation originated, as well as the *evidence score*. The evidence score is between 1 and 4 and refers to the number of mapping sources (Approximate Term API, Averbis text analysis pipeline, Historical mapping, RxNorm embedding nearest neighbor search) that produced the same mapping. Given this information, the prompt requests `GPT4o` to output a score between 0 and 10, where higher scores should denote a higher likelihood of mapping correctness.

### Prediction of human review using OpenAI Embeddings and LightGBM

A key component of our pipeline is a probabilistic classifier, that assesses whether a mapping of a local description to a RxCUI description is correct. For machine learning, we create a feature vector by stacking two 128 dimensional embeddings, one of the local description and one of the RxCUI description, generated using an OpenAI-based `text-embedding-small` model, and combine it with 7 additional features: 4 binary indicator variables that represent which of the 4 methods in the voting ensemble agreed on the mapping, the `GPT4o`-based pre-review score, the ingredient count (the number of ingredient mappings that were produced for a given local description), as well as a categorical variable of the country from which the medication description originated. The machine learning labels were provided by human review of a set of generated mappings (*n* = 2551 from 2 HCOs in Brazil, *n* = 9621 from 2 HCO in the United States, each evaluated by two Medical Informatics experts from Brazil, and the USA, respectively), assigning a label of 1 if the RxCUI mapping is correct, and 0 if it is incorrect. Finally a LightGBM-based binary classifier was trained to return the probability that a tentative mapping is correct. These probabilities were post-hoc calibrated using isotonic regression. We consider RxNorm mappings with a predicted probability of 1 to be correct in the final output of our pipeline, because we wanted to maximize precision of the mappings. The data that was used for the training is sourced from different HCOs than the 6 HCOs that were used in the evaluation experiments, so no data leakage into the test set could occur.

### Evaluation metrics

We define recall as the proportion of local medication codes which have a correct RxNorm ingredient (IN) CUI mapping in the output of the pipeline. If `MIN` ingredient codes are in the gold standard (evaluation on the public data set), the definition of recall requires all related `IN` codes to be produced. Precision is defined as the proportion of generated mappings that are correct. The F1-score is the harmonic mean of the recall and precision.

## 4 Results

### Performance in public Med-UCD data set from France

To evaluate the performance of our pipeline, we compare its output with the Med-UCD (French Unite Commune de Dispensation) data set, as preprocessed by Rouhizadeh et al. [14]. Med-UCD contains local medication descriptions that are assigned to the correct RxNorm ingredient code. If the term type of this code is `MIN` we require the models to either produce the exact `MIN` code or all of the related `IN` RxCUIs, for defining the evaluation metric recall. 10 example descriptions from the data set are displayed in Table 3. We compare against the two best performing models benchmarked by Rouhizadeh et al. [14] (Table 4). To achieve a fair comparison between all methods, we exclude samples in the data set with an exact match in a lookup database, such as UMLS for the baselines or RxNorm description/Historical DB for `RxEmbed`. This yielded 2,612 unique evaluation terms.

**Table 3:** Example of 10 pairs of local medication description, RxNorm codes and their descriptions randomly sampled from the French Unite Commune de Dispensation (Med-UCD) data set (n=2,612)

| Medication description | RxNorm code | RxNorm description |
| --- | --- | --- |
| zolmitriptan, zomig 2,5mg cpr nsfp | 135775 | zolmitriptan |
| metformine, metformine isd 850mg cpr | 6809 | metformin |
| betamethasone, celestene 4 mg sol inj | 1514 | betamethasone |
| abacavir/lam myp600/300mg cp | 614534 | abacavir / lamivudine |
| silodosine, urorec 4 mg gelule | 720825 | silodosin |
| iode, ultravist 18.5g sol inj | 27781 | iopromide |
| fluticasone, avamys 27,5 microg/pulv susp nasale | 705022 | fluticasone furoate |
| potassium, potassium chl mrm 15% inj 10ml nsfp | 8591 | potassium chloride |
| memantine, memantine bga 20 mg cpr | 6719 | memantine |
| losartan, cozaar 2,5 mg/ml pdr et sol buv | 52175 | losartan |

**Table 4:** Performance of RxEmbed in the French Unite Commune de Dispensation (Med-UCD) data set for generating RxNorm ingredient mappings, compared with two baselines, as described by Rouhizadeh et al. [14]. Proposed mappings were evaluated by comparing against the gold standard RxNorm mappings contained in the data set. Sample size refers to the number of local descriptions, for recall, and the number of mappings, for precision, that were assessed.

| Model | F1-score | Recall (Sample size) | Precision (Sample size) |
| --- | --- | --- | --- |
| RxEmbed (ours) | **90.9 %** | **88.9 %** (n=2612) | **92.9 %** (n=2566) |
| e5 (Baseline [14]) | 83.3 % | 83.3 % (n=2612) | 83.3 % (n=2612) |
| BioMistral (Baseline [14]) | 84.9 % | 84.9 % (n=2612) | 84.9 % (n=2612) |

### Performance on TriNetX HCOs from the United States

From TriNetX’s network of HCOs in the US, we have selected 3 HCOs, each from a different census region of the country. They were selected because their medication data was not represented by a standard terminology where we could perform a code-to-code mapping, and required to be mapped by drug name. Each organization had unique subjective characteristics in their medication descriptions, and differed regarding their mix of brand and generic medications, number of tokens, and string length. Example descriptions from the data set are displayed in Table 6. We applied `RxEmbed` to the local descriptions originating from each HCO. The resulting mappings were then reviewed by an expert in medical informatics and pharmacology from the United States, and then evaluated for recall and precision, as defined above (Table 5).

**Table 5:**
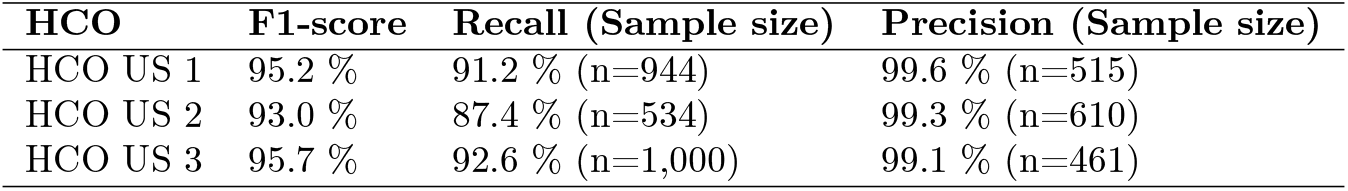
Performance of RxEmbed in the TriNetX data set of medication description/codes gathered from 3 healthcare organizations (HCOs) in the United States. The mappings were manually reviewed to assess recall/precision. Sample size refers to the number of local descriptions, for recall, and the number of mappings, for precision, that were assessed.

**Table 6:**
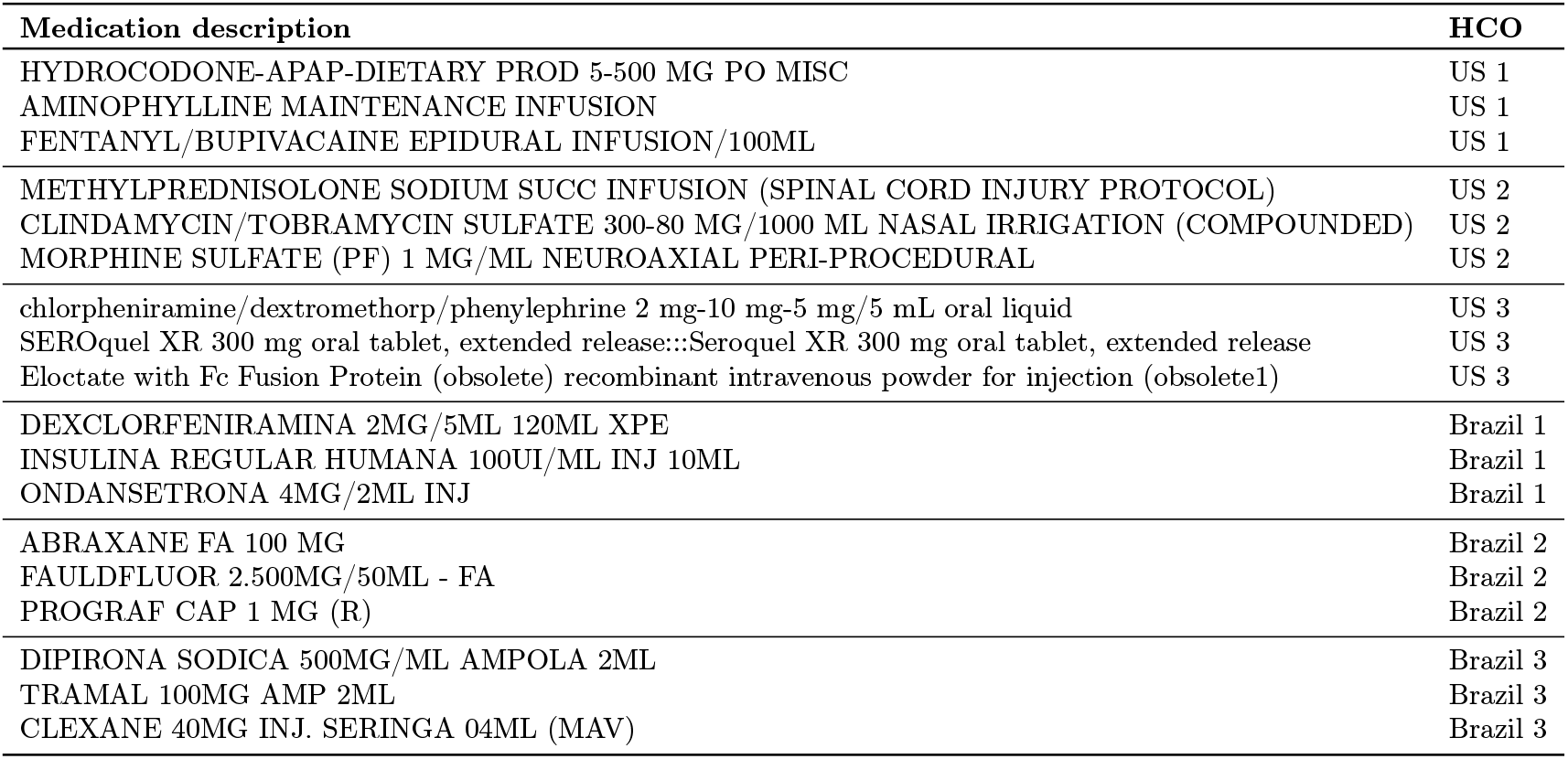
Example of local medication descriptions from the 3 selected healthcare organizations (HCOs) in TriNetX’s network in the US, and the 3 selected HCOs in TriNetX’s network in Brazil.

### Performance on TriNetX HCOs from Brazil

From TriNetX’s network of HCOs in Brazil, we have selected 3 HCOs, a general cancer center from the Northeast Region, a general hospital from the Southeast region, and a Hospital Network/Chain over 10 units. The nature of the local medication descriptions differed slightly. Whereas HCO Brazil 1 had mainly drug names and oncology compounds, HCO Brazil 2 used mostly product/brand names. Finally, HCO Brazil 3 contained a mixture of the above. Example descriptions from the data set are displayed in Table 6. We applied `RxEmbed` to local descriptions originating from each HCO. The resulting mappings were then reviewed by an expert in medical informatics and pharmacology from Brazil and evaluated in terms of recall and precision, as defined above (Table 7).

**Table 7:** Performance of RxEmbed in the TriNetX data set of medication description/codes gathered from 3 healthcare organizations (HCOs) in Brazil. The mappings were manually reviewed to assess recall/precision. Sample size refers to the number of local descriptions, for recall, and the number of mappings, for precision, that were assessed.

| HCO | F1-score | Recall (Sample size) | Precision (Sample size) |
| --- | --- | --- | --- |
| HCO Brazil 1 | 95.3 % | 91.5 % (n=961) | 99.5 % (n=402) |
| HCO Brazil 2 | 91.9 % | 85.0 % (n=1245) | 100.0 % (n=303) |
| HCO Brazil 3 | 91.1 % | 84.0 % (n=2000) | 99.5 % (n=428) |

## 5 Discussion

We built and evaluated a novel LLM-based medication mapping pipeline, which we call `RxEmbed`, that binds local medication descriptions from EHR systems to RxNorm ingredient codes. Such an approach has to be used when the coding systems used to represent medication data are not directly mappable to standard terminologies such as RxNorm by code. Harmonizing all medication data to a uniform standard, in this case RxNorm, is crucial for a seamless operation of a global federated network of EHR data such as TriNetX.

Our pipeline supports entity extraction from medication descriptions, binding to RxNorm ingredient codes, fulfilling a critical operational need for scalable, automated mapping review. This ML-based automation is the novel component of the pipeline. It reduces the manual effort required to review the output of a mapping pipeline, which can be very resource-intensive when mapping tens of thousands of distinct medication concepts.

When comparing the performance of the pipeline between medication data in English (USA) and Portuguese (Brazil), it is apparent that while recall is quite high in all 6 evaluated sites, the recall for US sites (87-93 %) is slightly higher than the recall for Brazilian sites (84-92 %). This could be due to various factors: (1) translation from Portuguese to English has to be performed, which could introduce errors; (2) increased difficulty when recognizing Brazilian brand names, both in large language models, and for NLP methods such as Approximate Term API and Averbis’ HealthDiscovery; and (3) the target terminology RxNorm being US-centric, which favors the use of the pipeline for drugs on US market. We expect that for countries outside of the US, pipeline performance could be increased further by introducing OMOP RxNorm Extensions [13] as a mapping target besides RxNorm. The precision, as evaluated by hand-reviewing generated mappings for internal TriNetX data, was very close to 100 % for our pipeline, but was slightly reduced (93 %) for the French Med-UCD data set. This is likely due in part to characteristics of the Med-UCD dataset, which, as we noted, may occasionally miss appropriate IN codes in the gold standard, since each description is linked to only one RxCUI.

### Limitations

This work has some limitations. Certain experiments were performed with proprietary TriNetX data from US and Brazil. However, we also obtained results on the mapping of the public data set Med-UCD from France, which allowed us to compare the recall/precision performance of our pipeline to other models and approaches. Ultimately, performance was assessed for data from three different countries in three different languages. In future work, the pipeline should also be assessed for data from other countries, including, for example, Japan and Taiwan, where obtaining the correct, information-preserving translation of native descriptions is challenging. Future studies should be performed to further validate this methodology, using publicly available data sets containing medication descriptions, and alternative LLM models such as LLAMA-3 or Mistral. `RxEmbed` readily supports substituting `GPT4o` with any other model that supports a chat completion interface.

## 6 Conclusion

We built and evaluated a LLM-based medication mapping pipeline, that binds local medication descriptions from EHR systems to RxNorm ingredient codes. The high precision of the pipeline output implies very limited need for human review of the generated mappings.

## Data availability

The TriNetX evaluation data are available from TriNetX, LLC but third-party restrictions apply to the availability of these data. The data were used under license for this study with restrictions that do not allow for the data to be redistributed or made publicly available. However, for accredited researchers, the TriNetX data is available for licensing at TriNetX, LLC. Data access may require a data sharing agreement and may incur data access fees.

## Consent to participate

This retrospective study is exempt from informed consent. The data reviewed is a secondary analysis of existing data, does not involve intervention or interaction with human subjects, and is de-identified per the de-identification standard defined in Section §164.514(a) of the HIPAA Privacy Rule. The process by which the data is de-identified is attested to through a formal determination by a qualified expert as defined in Section §164.514(b)(1) of the HIPAA Privacy Rule. This formal determination by a qualified expert refreshed on December 2020.

## A Tradeoff between precision and recall for different classification thresholds

We have observed empirically that the ‘predict human-review model’ produces an almost bi-modal distribution of scores in its output, after calibration with isotonic regression. The resulting distribution of scores is shown in Figure 2

**Figure 2.**
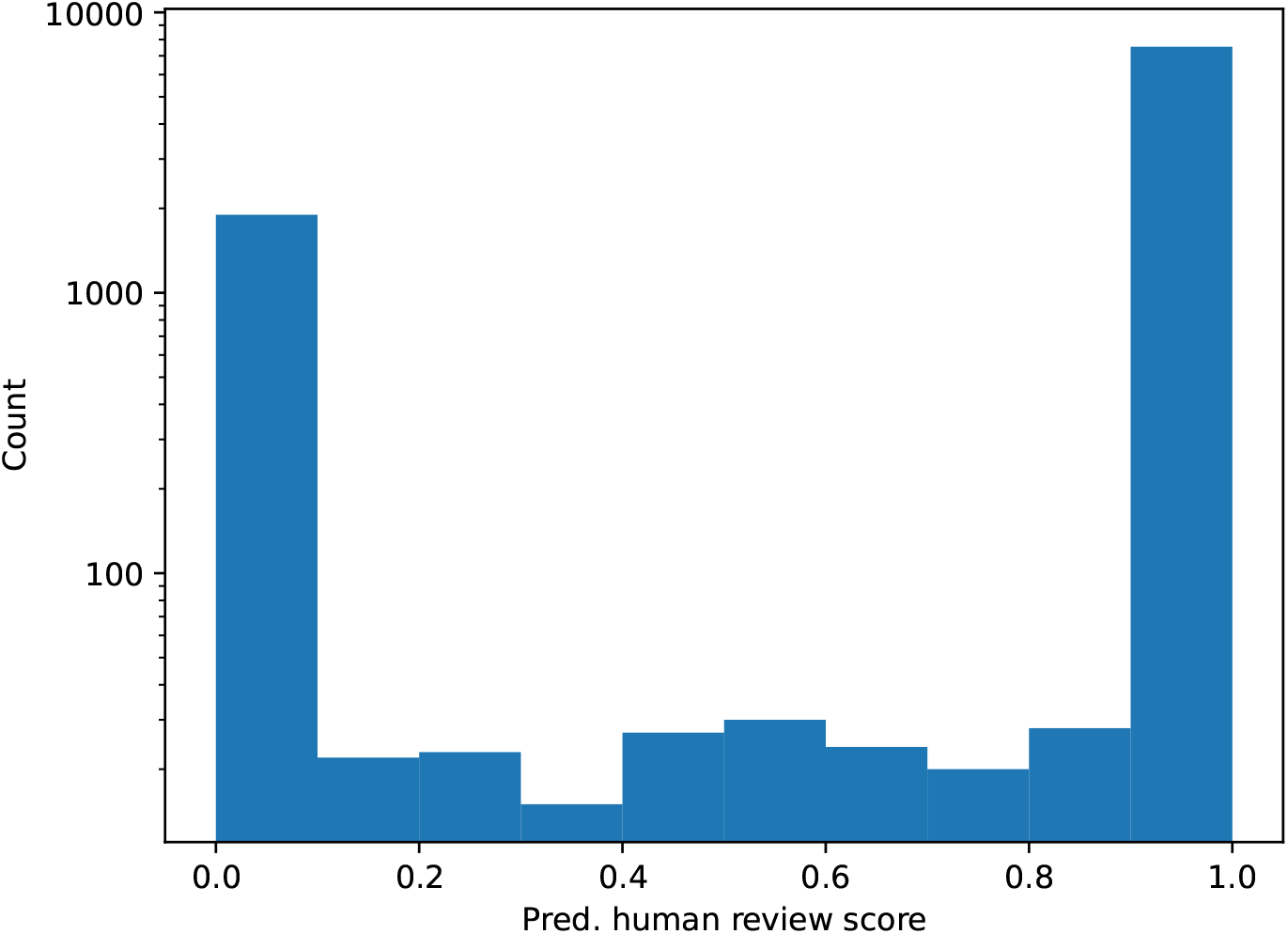
Distribution of scores of the predict human review model. The y-axis is an logarithmic scale to allow comparison of the two majority bins and the bins for intermediate scores not exactly equal to 0 or 1.

Moreover we analyzed the recalls and precisions that arise if a different threshold from 1 is chosen, which is used for the main experimental results. We observe that the effect of the decision threshold is relatively low, due to the small number of intermediate scores between 0 and 1 (Table 8). Note that even at a decision threshold of 0 the recall is not necessarily 100 % because not all required mappings are produced by the pipeline

**Table 8:** Performance of RxEmbed in the public dataset Med-UCD for generating RxNorm ingredient mappings, at different decision thresholds to explore the tradeoff between precision and recall of the generated pipeline output in the different variants. Results for for the best-performing configuration for each metric are shown in bold-face.

| Decision threshold | F1-score | Recall | Precision |
| --- | --- | --- | --- |
| 1 | **90.9 %** | 88.9 % | **92.9 %** |
| 0.9 | 90.7 % | 89.0 % | 92.5 % |
| 0.8 | 90.6 % | 89.1 % | 92.2 % |
| 0.7 | 90.5 % | 89.1 % | 92.0 % |
| 0.6 | 90.4 % | 89.1 % | 91.8 % |
| 0.5 | 90.3 % | 89.2 % | 91.4 % |
| 0.4 | 90.2 % | 89.2 % | 91.2 % |
| 0.3 | 90.2 % | 89.4 % | 91.1 % |
| 0.2 | 90.2 % | 89.4 % | 91.0 % |
| 0.1 | 90.1 % | 89.4 % | 90.9 % |
| 0.0 | 82.7 % | **89.6 %** | 76.8 % |

